# GEOGRAPHIC DOMAIN SHIFT PRECIPITATES DIVERGENT FAILURE MODES IN DEEP LEARNING–BASED TUBERCULOSIS SCREENING: A MULTI-NATIONAL EXTERNAL VALIDATION STUDY

**DOI:** 10.64898/2026.01.17.26344327

**Authors:** Ibrahim Ibrahim Shuaibu, Muhammad Ayan Khan, Diyaa Alkhamis, Anas Alkhamis

## Abstract

**Background:** Deep learning algorithms for tuberculosis (TB) screening frequently achieve radiologist-level performance during internal evaluation, yet their reliability often degrades when deployed to populations differing from the training domain. Such degradation is clinically consequential for screening tools, where the World Health Organization (WHO) emphasizes high sensitivity to minimize missed infectious cases.

**Methods:** A DenseNet-121 convolutional neural network was trained using transfer learning on the Shenzhen chest X-ray dataset (China; total n=662). To prevent anatomically implausible augmentation, horizontal flipping was excluded during training. The model was trained in two stages (head training followed by fine-tuning) and evaluated on: (i) an internal test set from China, (ii) an external balanced cohort from Montgomery County (USA; n=138), and (iii) an external TB-positive cohort from India (n=155). The India dataset served as a sensitivity stress test; specificity and ROC-AUC were not computed for this cohort due to the absence of negative controls. Model attention was explored using Grad-CAM.

**Results:** Internal validation yielded an Area Under the Curve (AUC) of 0.889 and accuracy of 85.6%. External testing revealed divergent failure modes. On the USA cohort, sensitivity was high (94.8%) but specificity decreased significantly (43.7%), indicating false-positive inflation. Conversely, on the India TB-only cohort, sensitivity collapsed to 52.3%, implying that 47.7% of confirmed TB cases were missed under domain shift. All metrics are reported as point estimates

**Conclusion:** Geographic domain shift produced non-uniform degradation false-positive surges in a low-burden setting and sensitivity collapse in a high-burden setting. These findings highlight the safety risks of deploying single-source TB screening AI without local validation and calibration.

## 1. Introduction

Tuberculosis (TB) remains the leading cause of death from a single infectious agent worldwide, disproportionately affecting low- and middle-income countries [1]. Chest radiography (CXR) is the primary triage tool for pulmonary TB; however, its diagnostic utility is constrained by significant inter-reader variability, high workload, and a shortage of expert radiologists in high-burden regions [2, 3]. Consequently, the World Health Organization (WHO) has recommended Computer-Aided Detection (CAD) software as an alternative to human interpretation for screening and triage in individuals aged 15 years and older [4].

While deep learning models, particularly Convolutional Neural Networks (CNNs), have demonstrated high performance in controlled studies [5, 6], a critical barrier to clinical safety is “domain shift” the degradation of model performance when applied to data distributions different from the training set [7]. Variations in patient demographics, disease prevalence, and image acquisition parameters (e.g., tube voltage, scanner manufacturer) can introduce site-specific artifacts that models may exploit as “shortcuts” rather than learning robust pathological features [8, 9].

The clinical consequences of domain shift are not uniform. A model may fail by missing active cases (false negatives), thereby perpetuating transmission, or by flagging healthy individuals (false positives), overwhelming healthcare systems with unnecessary confirmatory molecular testing [10]. Despite this, few studies rigorously “stress-test” models across disparate geographic borders to characterize these specific failure modes [11].

This study evaluates the cross-border generalizability of a DenseNet-121 model trained exclusively on data from China. By testing the model on unseen cohorts from the USA and India, we test the hypothesis that geographic domain shift precipitates divergent clinical failure modes, necessitating population-specific calibration.

## 2. Materials and Methods

### Study Design and Data Sources

This retrospective external validation study utilized three geographically distinct datasets (Table 1). The study design prioritized the prevention of data leakage and the explicit handling of single-class testing for the India cohort.

**Table 1:**
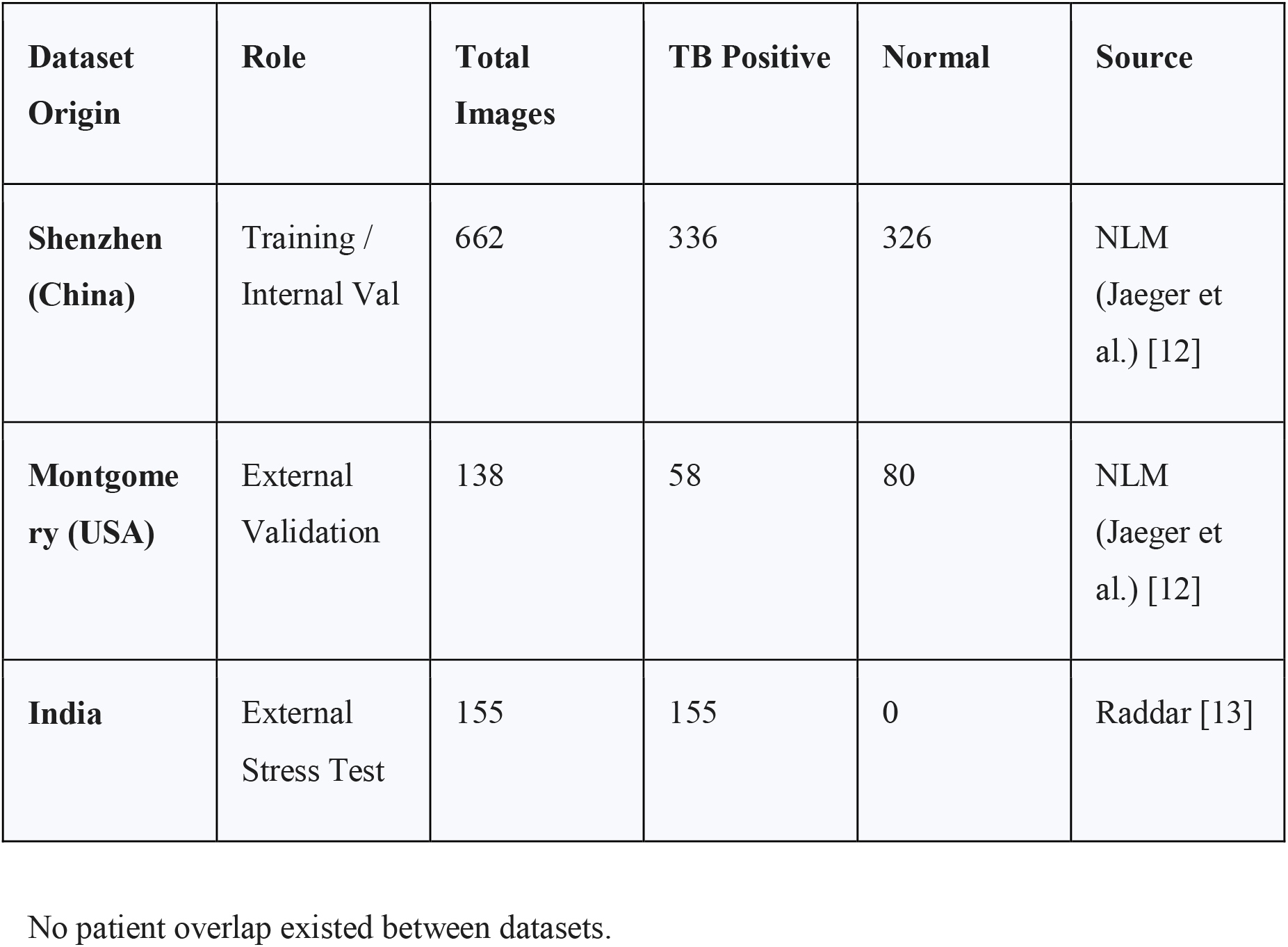
Characteristics of the datasets used in this study.

| Dataset Origin | Role | Total Images | TB Positive | Normal | Source |
| --- | --- | --- | --- | --- | --- |
| Shenzhen (China) | Training / Internal Val | 662 | 336 | 326 | NLM (Jaeger et al.) [12] |
| Montgomery (USA) | External Validation | 138 | 58 | 80 | NLM (Jaeger et al.) [12] |
| India | External Stress Test | 155 | 155 | 0 | Raddar [13] |

**Table 1.** Diagnostic Performance Metrics of the Proposed Model.

| Performance Metric | Value (%) |
| --- | --- |
| Accuracy | 85.6 |
| Sensitivity | 77.6 |
| Specificity | 49.2 |
| AUC (ROC) | 0.889 |

**Table 2:** Summary of Model Performance.

| Dataset | Split Type | Accuracy | Sensitivity | Specificity | AUC |
| --- | --- | --- | --- | --- | --- |
| China<br>(Shenzhen) | Internal Val | 85.6% | 77.6% | 49.2% | 0.889 |
| **USA**<br>(Montgomery) | External Test | 65.2% | **94.8%** | **43.7%** | 0.822 |
| **India**<br>(Raddar) | Stress Test | 52.3%* | **52.3%** | N/A | N/A |

#### Training Source (China)

The Shenzhen Hospital X-ray Set [12] was selected for model development due to its balanced class distribution and high image quality.

#### External Test A (USA)

The Montgomery County X-ray Set [12] served as a balanced external validation set representing a low-burden, high-resource setting.

#### External Test B (India)

A cohort of 155 radiologist-confirmed TB-positive images from India [13] was utilized. As this dataset contained no negative controls, it was employed strictly as a “Sensitivity Stress Test.”

No patient overlap existed between datasets.

### Pre-processing and Augmentation

Radiographs were resized to 224 × 224 pixels. Input normalization followed the standard DenseNet pre-processing protocol (channel-wise mean subtraction and scaling) to align with ImageNet weights.

Data augmentation was applied during training to improve robustness, including random rotations (pm 10^{circ}) and zooming (0.95x - 1.05x). Crucially, **horizontal flipping was explicitly excluded**. Unlike natural images, chest radiographs possess anatomical laterality (e.g., the cardiac apex is left-sided). Flipping images can create non-physiological artifacts (dextrocardia) that may confound the model’s ability to localize pathology [14]. Images were scaled to [0,1] prior to normalization.

#### Model Architecture

We employed a DenseNet-121 architecture [15], initialized with weights pre-trained on ImageNet [16]. DenseNet was selected for its feature reuse capability, which is particularly effective for medical imaging tasks with limited data. The architecture utilizes dense connections between layers to improve gradient flow. “A sigmoid output was used to enable binary probabilistic classification.”

The classification head was modified to include a Global Average Pooling layer, a dense layer (512 units, ReLU activation), a dropout layer (rate 0.5) to mitigate overfitting [17], and a final sigmoid output unit.

### Training Protocol

To prevent catastrophic forgetting of pre-learned features, training proceeded in two stages:

#### Head Training

The convolutional base was frozen, and the classification head was trained for 8 epochs (Adam optimizer, learning rate 1 × 10^{−4}).

#### Fine-Tuning

The top 30 layers of the base were unfrozen, and the model was fine-tuned for 6 epochs with a reduced learning rate (1 × 10^{−5}) [18].

### Evaluation Metrics

Performance was assessed using Accuracy, Sensitivity (Recall), Specificity, and Area Under the Receiver Operating Characteristic Curve (AUC).

For the **India dataset (TB-only)**, Specificity and AUC are statistically undefined (division by zero). Therefore, results for India are reported solely as Sensitivity and False Negative Rate. Model interpretability was assessed using Gradient-weighted Class Activation Mapping (Grad-CAM) [19] to visualize regions of interest.

## 3. Results

### Internal Validation (China)

On the internal hold-out set from Shenzhen, the model demonstrated robust discrimination.

The confusion matrix indicated a balanced performance profile, consistent with effective learning of the source domain features.

### External Validation (USA): The “False Positive Surge”

Upon evaluation of the US cohort, the model exhibited a shift in operating characteristics. Sensitivity increased to 94.8% (55/58 cases detected), but specificity declined markedly to 43.7% (35/80 healthy cases correctly identified).

This indicates a “high alert” behavior where the model essentially over-calls pathology, resulting in 45 false positives. While the AUC remained respectable (0.822), the low specificity would significantly burden clinical workflows.

### External Validation (India): The “Sensitivity Collapse”

Testing on the India cohort revealed a critical safety failure. Despite the dataset consisting entirely of confirmed TB cases, the model identified only 81 out of 155 cases.

This performance represents a collapse in diagnostic capability, where the model failed to recognize nearly half of the active cases in this high-burden population. This failure is visualized in the sensitivity comparison in Figure 3. These results do not represent diagnostic accuracy but stress-test sensitivity under domain shift.

**Figure 1:**
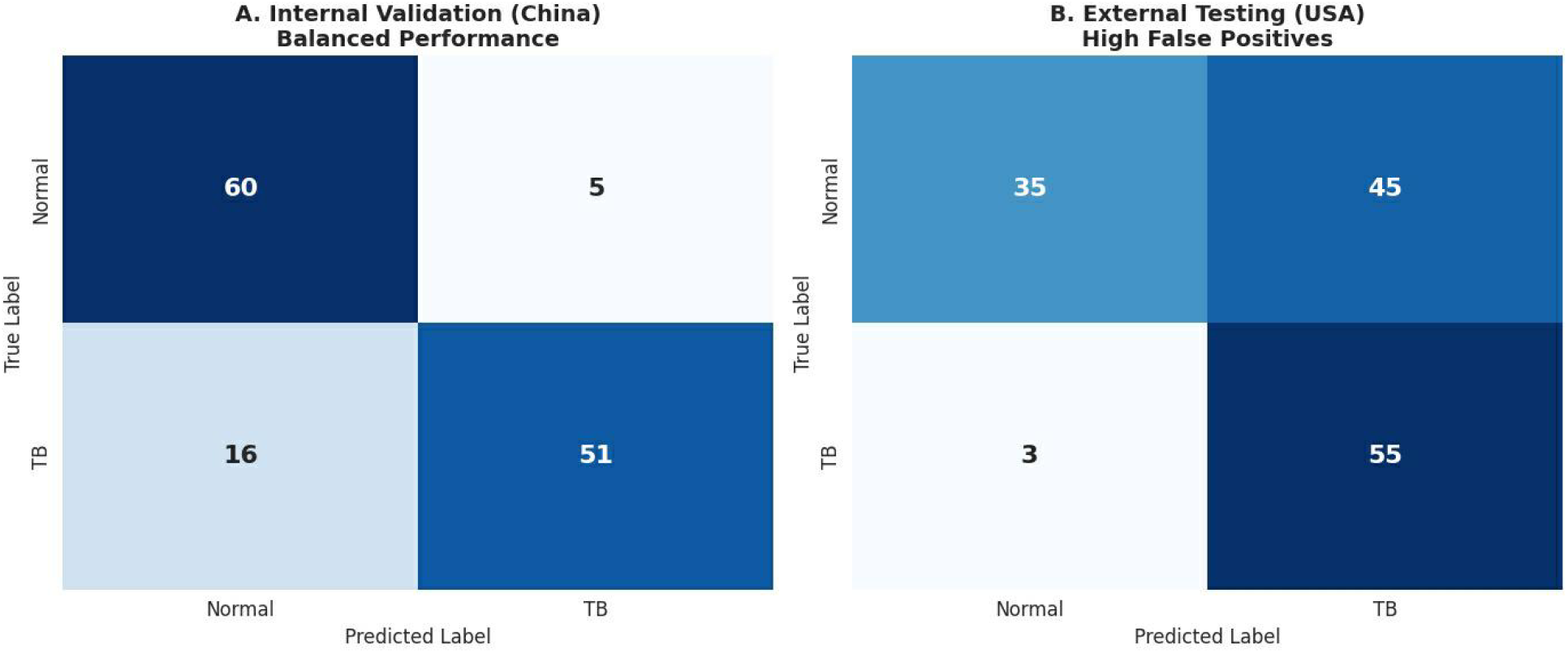
Confusion Matrices comparing China (A) and USA (B)

**Figure 2:**
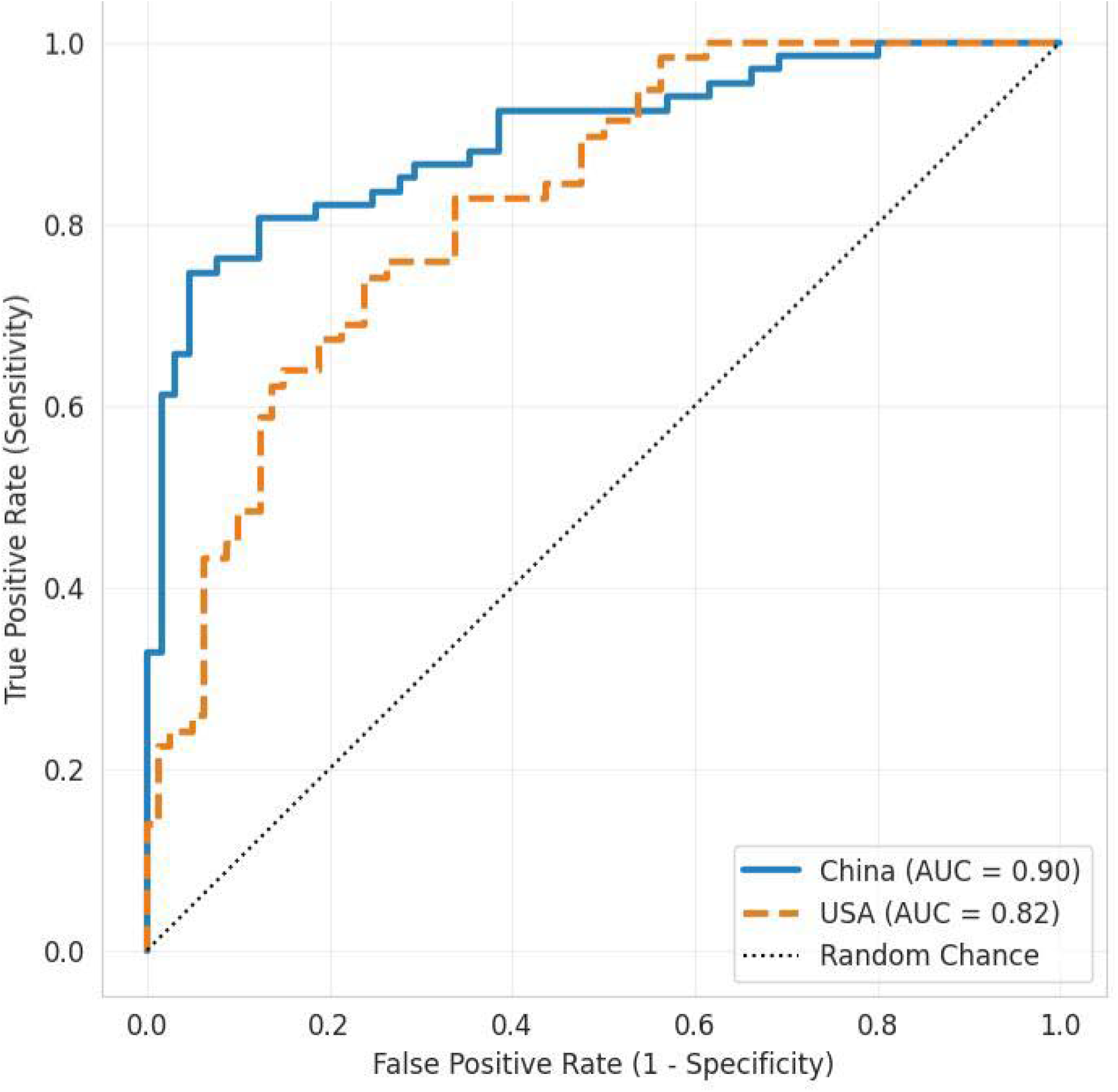
ROC Curves for China and USA.

**Figure 3:**
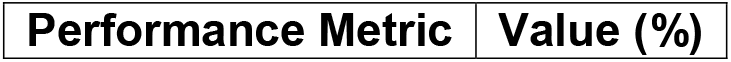
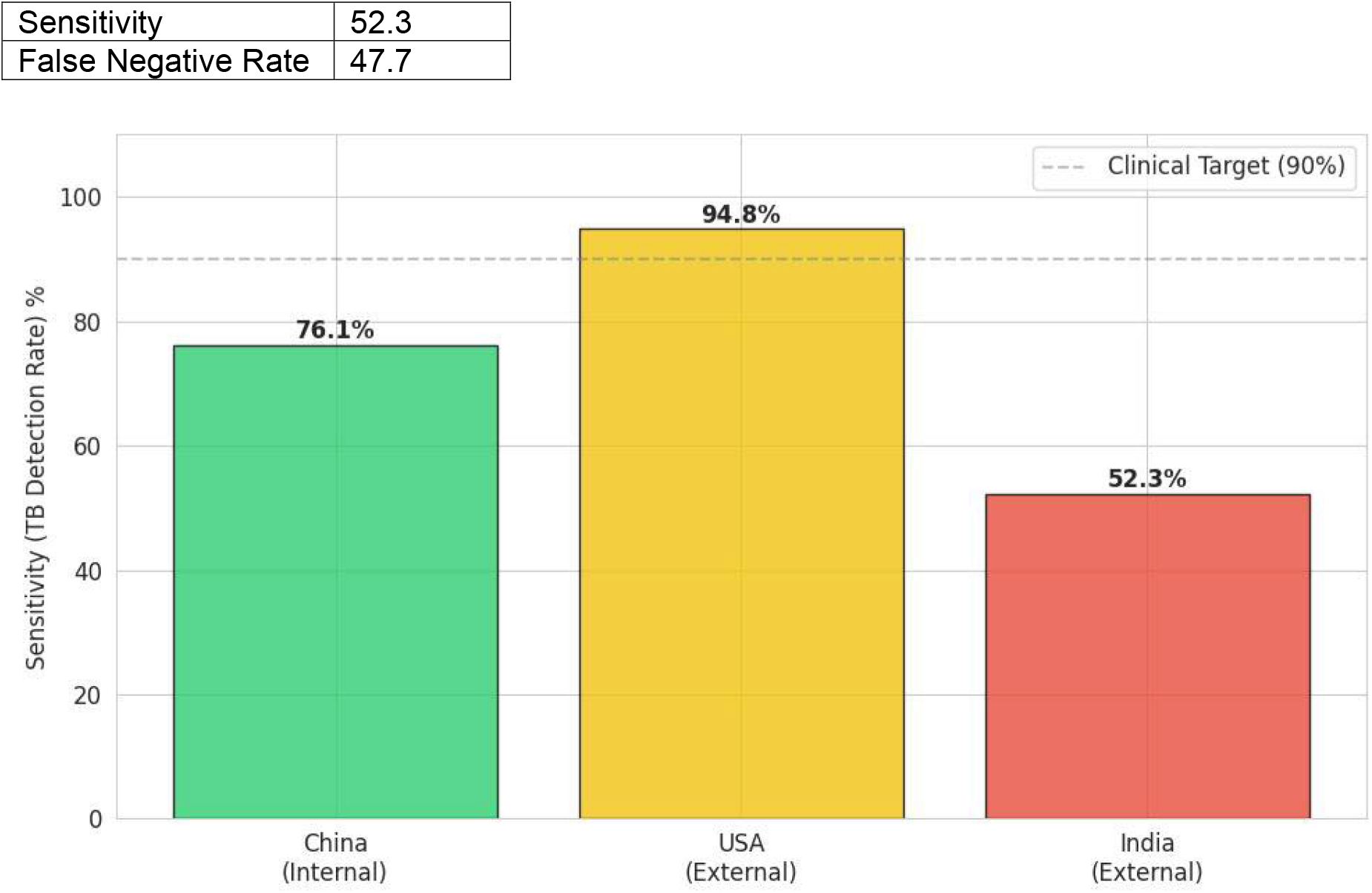
Bar Chart showing Sensitivity Collapse across datasets.

### Explainability

Grad-CAM heatmaps (**Figure 4**) for correctly classified India cases showed activation in the lung fields, confirming that the model utilizes anatomical features when it succeeds. However, the high false-negative rate suggests that for the missed 47.7% of cases, the pathological features (e.g., cavitation, consolidation) in the Indian cohort did not trigger the feature detectors learned from the Chinese data.

**Figure 4:**
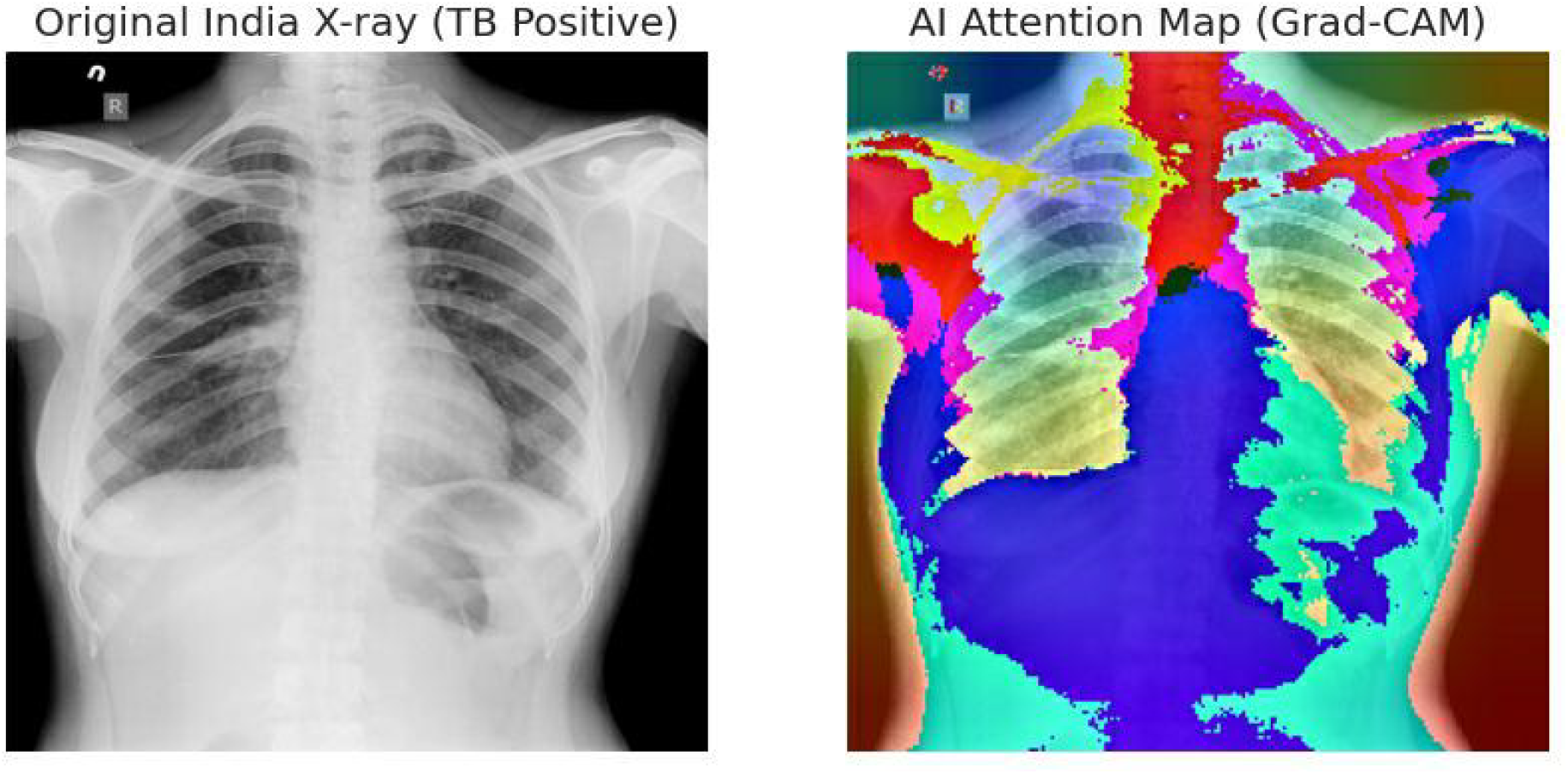
Grad-CAM Heatmap overlay.

## 4. Discussion

This study provides empirical evidence that deep learning models for TB screening lack inherent cross-border robustness. We identified two distinct, geography-dependent failure modes: a “False Positive Surge” in the USA and a “Sensitivity Collapse” in India.

### Divergent Failure Modes

The degradation in the USA cohort manifested as reduced specificity. This phenomenon, often termed “domain shift,” likely results from differences in image acquisition [20]. American radiographs in the Montgomery set are historically noted for different contrast and brightness profiles compared to Shenzhen images. The model appears to have interpreted the site-specific “haze” or noise of the US scanner as pathological opacity, leading to high sensitivity but poor specificity. In practice, this leads to “alert fatigue,” where clinicians ignore AI warnings due to excessive false alarms [21].

Conversely, the failure in the Indian cohort is clinically more dangerous. Missing 47.7% of active cases renders the model unsafe for screening in this setting. TB presentation in India often involves advanced disease states, including extensive fibrosis or cavitation [22], which may differ morphologically from the cases in the training set. Furthermore, variations in patient positioning (AP vs. PA views) and chest wall adiposity may contribute to the loss of signal detection [23].

### Limitations

This study has limitations. The India dataset lacked healthy controls, preventing the calculation of specificity or AUC for that region. Additionally, we utilized a single architecture (DenseNet-121); while this is a standard benchmark, ensemble methods might offer improved robustness [24]. Finally, we did not apply unsupervised domain adaptation techniques, which could theoretically align feature distributions between countries [25].

## Conclusion

The generalizability of TB screening AI cannot be assumed. A model that is 85% accurate in China may fail to detect half the cases in India or falsely flag healthy patients in the USA. These findings confirma that local external validation is not merely a regulatory step but a clinical safety imperative. We recommend that future deployments incorporate local calibration thresholds to mitigate these divergent failure modes.

## Data Availability

All data produced in the present study are available upon reasonable request to the authors

## Declaration

The authors declare that no funds, grants, or other support were received during the preparation of this manuscript.

The author declares no conflicts of interest. Clinical trial number: not applicable

## Author Contributions

All authors contributed substantially to the conception and design of the study. The computational model was developed, implemented, and validated by the research team. Data analysis, simulation, and interpretation of results were performed collaboratively. All authors contributed to drafting the manuscript and critically revising it for important intellectual content. All authors approved the final version of the manuscript and agree to be accountable for all aspects of the work, ensuring its accuracy and integrity.

